# Evaluation of the US governors’ decision when to issue stay-at-home orders

**DOI:** 10.1101/2020.05.14.20093633

**Authors:** Benjamin Djulbegovic, David J. Weiss, Iztok Hozo

**Author notes:** Corresponding author: Benjamin Djulbegovic, MD, PhD, City of Hope, 1500 East Duarte Rd.,Duarte, CA. 91010.

## Abstract

**Objectives:** To evaluate if the US governors’ decision to issue the stay-at-home orders reflects the classic Weber-Fechner law of psychophysics, the amount by which a stimulus (such as number of cases or deaths) must increase in order to be noticed-the *“just noticeable difference”-*as a fraction of the intensity of that stimulus.

**Design:** A prospective observational study using data on the daily number of infected patients and deaths from the New York Times daily database.

**Setting:** 50 States and the District of Columbia

**Participants:** All individuals judged to be positive for the coronavirus or to have died from COVID19.

**Main outcome measures:** Number of people diagnosed with or died from COVID19.

**Results:** We found that the decision to issue the state-at-home order reflects the Weber-Fechner law of psychophysics. Both the number of infections (p=<0.0001; R^2^=0.79) and deaths (p<0.0001; R^2^=0.63) were highly statistically significantly associated with the decision to issue the stay-at-home orders. The results indicate that for each doubling of infections or deaths within their state, an additional four to six governors will issue the stay-at-home order. We also observed a clear dose-response relationship in the Cox model: the larger the number of cases, or deaths, the higher the probability that the stay-at-home order will be made. When the number of deaths reached 256 or the number of infected people was greater than 16,384, the probability of issuing a “stay-at-home” order was close to 100%.

**Conclusions:** When there are not clearly articulated rules to follow, decision-makers in times of crisis such as COVID19 resort to use of simple heuristics consistent with the Weber-Fechner law of psychophysics. The findings are important for the public to understand how their elected officials make important public health decisions.

*Strengths and limitations of this study:*

- The study was based on individual, publicly available patient data accessible to both researchers and policy-makers.
- The US governors seemed to be wired to act according to the Weber-Fechner law of psychophysics
- The Weber-Fechner function explained 60-80% of the variance in the governors’ decisions. Typically, explanatory factors in decision-making fields capture at best 30% of such variance.
- The accuracy of data and reporting practices could not be independently verified.
- We demonstrated an association but not a causal relation-we have identified the stimulus that underlies the decision, but we do not know why some governors acted more quickly than others.

## Introduction

As the COVID-19 pandemic spreads, keeping people apart has been hailed as the most important preventive measure currently available. In the U.S., the reluctance of the federal government to issue a national order has placed state governors in the position of having to decide when to escalate from vague wishes for social distancing to stay-at-home directives. What information guides that decision? We suggest there are two clearly identified, readily available signals that convey crisis magnitude, namely the numbers of cases (people reported as infected) and people pronounced dead of the disease. When those numbers increase sufficiently, action will be taken. The two numbers are correlated, of course, but we might expect each of these numbers to affect the governor’s decision to issue a stay-at-home order.

People vary in their sensitivity to stimulus differences. According to the classical Weber-Fechner law of psychophysics^1 2^, the amount by which a stimulus must increase in order to be noticed, the *“just noticeable difference’’* or JND, is a fraction of the intensity of that stimulus. For example, Ernst Weber, a 19^th^ century psychologist, found that people could not discriminate between 20.5 and 20.0 g weights but could usually discriminate between 21 and 20 g.^2^ When a series of baseline weights was 40, 60, 80, and 100 g, the *JND* was 2, 3, 4, and 5 g, respectively. That is, to appreciate the *differences* between weights (*JND*), the weight (i.e., stimulus) should increase by a constant percentage of the stimulus itself i.e., by at least 5% of the original weights in this example.^2^ Gustav Fechner, another 19^th^ century psychologist, proposed that the *“JND”* can be conceptualized as a unit of psychological intensity rather than physical intensity.^1-3^ Subsequently, the relationship between the intensity of a signal and how much more intense the signal needs to increase before a person can reliably tell that the signal had changed has become known as the *Weber-Fechner Law* of psychophysics. As expected for physiological systems, the law is valid within certain domains of stimulus-response ratios. Therefore, if we accept Fechner’s assumption that *JNDs* are psychologically equal in size, then there will be a logarithmic relation between stimulus intensity and perception. The Weber-Fechner law has been well documented across different physiological and psychological phenomena ^4^ - it describes responses in a variety of domains in which increasingly intense stimuli can be placed along a continuum, from the perception of loudness or brightness^1 2^ to the subjective value of money^5^ to the mental line for numbers^6^ and risky choices.^7^ In the present context, we anticipate that the perception of danger from the virus will depend upon the number of cases and/or deaths observed within a governor’s jurisdiction.

Our hypothesis is that in order to move an uncommitted governor to action, the additional number of infected people or deaths required will be logarithmically larger as more cases or deaths are reported. That is, the governors will not “notice” that conditions are sufficiently worse until an increasingly larger number of people are infected or die.

## Methods

### Data

We obtained the daily numbers of cases and deaths from the New York Times daily database [https://github.com/nytimes/covid-19-data]. We examined both numbers separately, cases and deaths, to see whether one signal was more clearly linked to the stay-at-home decision. The analyses were performed on data downloaded on April 12, 2020 from all 50 states and the District of Columbia (we regarded the mayor of that city as a governor in our analyses); by that time all but 7 states had issued stay-at-home orders. Four of the governors had issued stay-at-home orders before any deaths were recorded.

The data cannot be regarded as precise. The Times consider a case confirmed when it is reported by a federal, state, territorial or local government agencies. Confirmed cases are patients who test positive for the coronavirus. However, it must be acknowledged that the performance of diagnostic tests vary, limiting diagnostic accuracy.^8^ Deaths are more visible, but may be inaccurate because of incorrect attributions. Some states updated records on a daily basis, while others updated only after a certain number of cases or deaths had accumulated. In sum, the reported numbers may not be exact, but the numbers we worked with and the numbers the governors saw are the same.

Because the virus surfaced at different times across the country, in order to count the number of days between the signaling event and the stay-at-home order, we had to start a separate calendar for each state. We called that starting date t_0_. We suspected that setting t_0_ to the day when the first death was reported would be appropriate, because officials are more likely to trust reports of deaths, which are concrete events, than of cases, which depend on the infected person’s decision to seek medical attention. It also would be possible to set t_0_ on the basis of cases. We tried that out to see if it mattered, but rather than starting the count after the first case in the state, as customarily in these types of the analyses, we started it after the first 50 cases had been reported. Because both cases and deaths accumulated exponentially, the growth rate was similar for both signals and so we expect similar results whichever way the calendar is defined.

### Statistical analyses

We used two analytical approaches. First, we fitted the Weber-Fechner logarithmic function by regressing the log_2_ of cases and deaths, respectively against the daily counts. If the logarithmic notion is correct, these should appear as straight lines. The idea is that relatively few cases or deaths may be needed to inspire the first governor to issue an order, while governors who do so later will require increasingly stronger signals. In the case of the latest states, many more cases or deaths may be needed. Using the base 2 for the logarithmic analysis allows us to see how many doublings of the signal strength (which we defined as JND) are required to inspire the various governors to issue the stay-at-home orders. Because we were interested in learning how the states responded in sequence, we first ranked the states according to the order of declaring the stay-at-home order after each state had cases ≥50 or deaths ≥1. We then regressed the stay-at-home state rank list on log_2_(cases) or log_2_(deaths). Thus, the coefficient of the regression represents the number of additional states introducing stay-at-home order for each doubling of cases or deaths.

For the analysis using cases, we excluded the 7 states that had not issued stay-at-home orders by April 12, the date of our analysis, thereby leaving us with data for 44 states. For the analysis using deaths, we also excluded an additional 4 states whose governors had issued the stay-at-home order before any deaths had occurred. The rationale for the latter is that these governors were not using the signals as we had defined them.

Another way to approach the data is to track the changes in the probability of making the stay-at-home decision as days pass. To do this, we used Cox regression modeling. We expected that the probability of issuing the stay-at-home order would proportionately increase as the number of cases or deaths increases. For the Cox analysis we used data for all eligible states but censored those states that had not issued the order by the time of our analysis

Because the data were somewhat noisy, we also repeated the analyses using three-day moving averages for cases and deaths. We performed sensitivity analyses using number of cases or deaths per million, and population size per area of state (density) as the stimuli rather than actual number. The results were sufficiently similar that we elected to not present them.

All analyses were done in STATA statistical software.^9^

### Patient and public involvement

This analysis is based on publicly available, individual data. No patients or public were involved in the design, recruitment and conduct of this study.

### Ethics/IRB approval

Not required. The analysis is based on publicly available, anonymized data.

### Data availability statement

Data are available at:https://github.com/nytimes/covid-19-data. Statistical code will be shared after the paper is accepted for publication.

### Results

Fig 1a and 1b show that both the numbers of cases (p=<0.0001; R^2^=0.79) and deaths (p<0.0001; R^2^=0.63) are significantly associated with the decision to issue the stay-at-home orders order. The results indicate that for each doubling of infections or deaths within their state, an additional four to six governors will issue the stay-at-home order. The analysis of the residuals for the linear regression analysis indicated no deviation from normality, thus supporting a customary assumption of the analysis.

**Figure 1:**
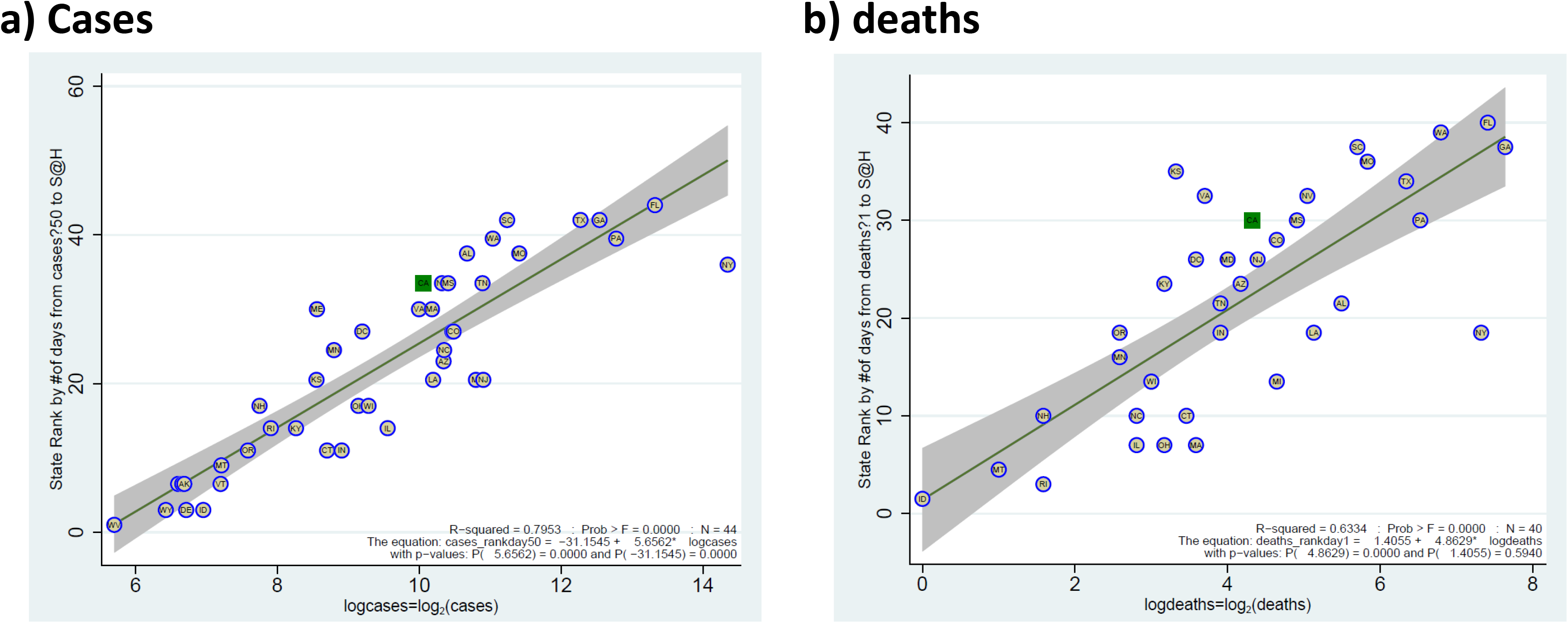
shows that the number of both cases (a) and deaths (b) is significantly associated with the decision to issue a “stay-at-home” order. For each doubling of cases beyond 50 or deaths beyond 1,it takes an additional 4-6 states to issue stay-at-home orders.

Our decision regarding t_0_, when to start each state’s calendar, was somewhat arbitrary. To see whether it mattered, we performed an additional set of exploratory analyses. In these, we started each state’s calendar after a particular number of cases or deaths were recorded. We tried fitting the logarithmic function to n’s from 1 to 100 for cases and from 1 to 20 for deaths. In general, as one would expect, better fit was observed when we started t_0_ at higher numbers of cases or deaths, but at the expense of fewer data points. For example, R^2^ increased to about 80% when we started at ≥10 deaths or ≥100 cases but was based on data from fewer states - 28 and 42, respectively.

The Cox regression curves in Fig 2a and 2b show that both number of cases and deaths were significantly associated with the probability of declaring the state-at-home order [hazard ratio (HR)=1.36 (p<0.045) for cases and HR=1.68 (p<0.00001) for deaths]. The adequacy of the analysis was confirmed by the non-significant result from Schoenfeld’s test for the proportional-hazard assumption, indicating the non-violation of the proportionality of effects of increasing number of cases or deaths on the probability of issuing state-at-home order.

**Figure 2.**
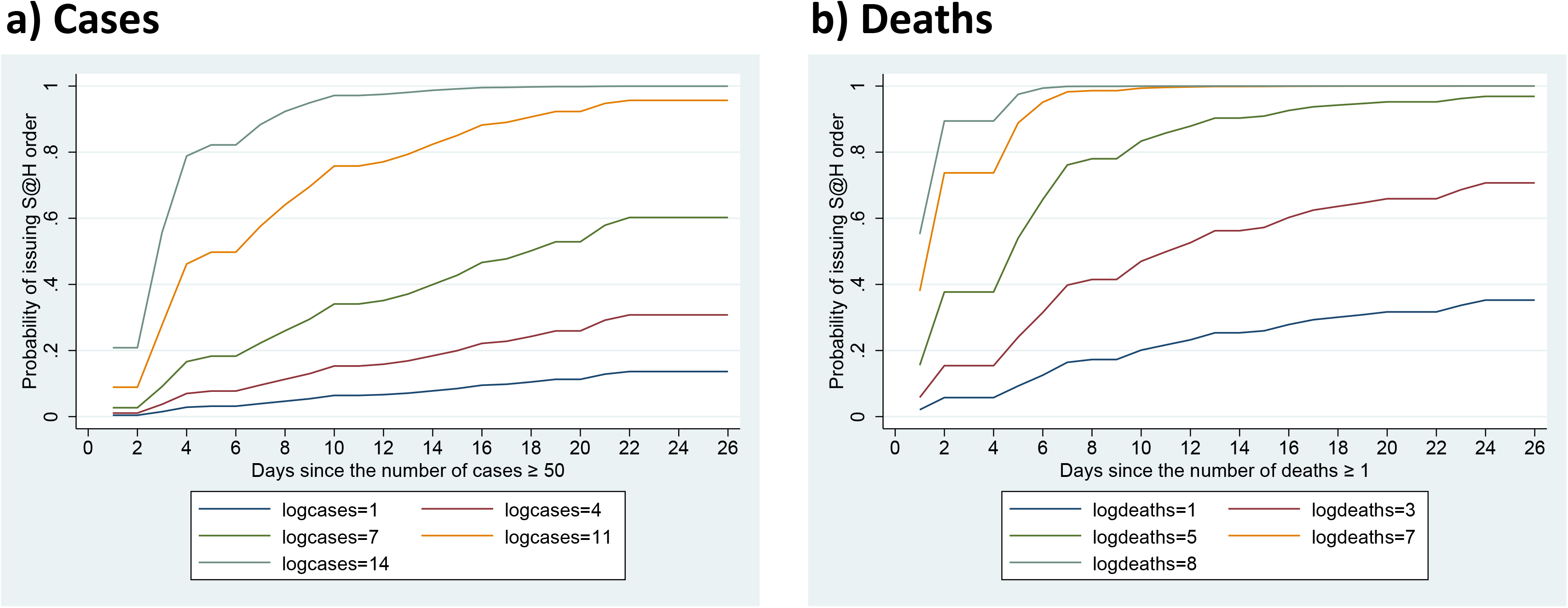
Cox regression analysis shows that the probability of issuing a stay-at-home order proportionally increases as the number of cases (a) and deaths (b) increases (the numbers expressed as an increase in log_2_ of cases or deaths are represented by different color lines on the graphs) [To convert log_2_(cases or deaths), raise the exponent to 2. For example, the largest number of cases in our database was log_2_(14) which converts into 2^14^= 16384; the largest number of deaths was log_2_(8) which translates into 2^8^= 256 deaths]. Abbreviations: S@H:stay-at-home order

We observed a clear dose-response relationship: the larger the number of cases, or deaths, the higher probability that the stay-at-home order will be made. The probability of issuing stay-at-home order was nearly 100% if the number of deaths exceeded log_2_(8) [i.e., 2^8^= 256 deaths]. For cases, this near-certainty occurs within 2 weeks of t_0_ when the number of cases exceeded log_2_(14) [i.e., 2^14^= 16,384 infected people]. The maximum number of cases or deaths for the states without stay-at-home orders were 2,221 and 94, respectively.

### Discussion

During the COVID19 crisis, one commentator remarked “we are all getting tired looking at the exponential graphs”, wondering how to act on the information shown in those graphs. From a theoretical perspective, we found that the decision to issue the state-at-home order reflects the classical psychophysical Weber-Fechner law of psychophysics. The signals – cases and deaths - are strongly correlated with the decision to announce the stay-at-home order. Here, for each doubling of the number of cases beyond 50 or deaths beyond 1, additional four to six governors will issue the stay-at-home order. When the signal is truly strong - more than about 250 deaths or 16,000 cases in our analysis-the probability of issuing state-at-home order approaches near-certainty.

Given the noise introduced by differences between states in terms of population density, political persuasion, and media coverage, this appears to be a remarkably simple result. The percentages of variance accounted for are quite high (60-80%) by industry standards, The typical R^2^ for a one-variable predictor in judgment research is between 5% to 30%. This appears to be another instance of a familiar phenomenon; when there are not clearly articulated rules to follow, people rely upon simple heuristics^10^ of which they need not be aware. These powerful rule-of-thumb, decision-making strategies can surprisingly be more accurate than complex statistical models, but sometimes can be disastrously wrong. ^10^ Here the heuristic is to wait until the day that the number of cases or deaths is striking. Indeed, it has been suggested that Weber-Fechner law can also operate as a heuristic –heuristic based on *the prominent numbers*efined as the powers of ten, their doubles, and their halves [e.g., 1,2, 5, 10, 20, 50, 100, 200…] approximates the Weber-Fechner law of psychophysics.^1 2 11 10^ For those governors who did not issue the state-at-home orders it is possible that they have not yet reached their personal Weber-Fechner heuristic threshold to act. Perhaps, crossing one of these psychologically important numbers-for example, more than 200 deaths may move the governors in that direction. There are, however, exceptions, as not all people employed this heuristic. The more proactive governors-the four who issued stay-at-home orders before any deaths were observed in their jurisdiction-seemed to have acted on the number of cases alone. And while the seven recalcitrant governors who did not issue orders may be waiting for an even stronger signal, it is also possible that their decisions are based on considerations of a completely different nature.

The major limitation of our approach is that it does not incorporate the kind of information - perhaps political stance, perhaps personality characteristics, perhaps availability of advice from colleagues or agencies - that might enable us to predict how many doublings in the numbers of deaths a particular governor needs to observe before taking action. Of course, our analysis demonstrated association between stimuli (cases, deaths) and the decision to issue a stay-at-home order, but not direct causation between the two. We have identified the stimulus that underlies the decision, but we do not know why some governors acted more quickly than others. As the governors seemed to be wired to act according to the Weber-Fechner law of psychophysics, we believe that the findings are important for the public to understand how their elected officials make important – life and death-public health decisions. This is ever more important as the states and countries are pondering their decisions how to re-open their economies.

## Data Availability

Data for the analysis reported in the paper are publicly available

https://github.com/nytimes/covid-19-data

## Acknowledgment

We want to thank Jerome Hoffman, MD and Dr. Amy Price for helpful comments on the earlier version of the paper.

## Authors’ contribution

Conceived idea, wrote the first draft, analysis (BD); revised draft, provided further input from the perspective of psychophysics (DJW); created a software program, analysis (IH)

